# Threshold Effects of Rehabilitation Intensity on Functional Recovery After Ischaemic Stroke: A Panel Threshold Regression Analysis of Australian Hospital Data

**DOI:** 10.64898/2026.03.11.26348201

**Authors:** Amaevia Lim, Priya Venkataraman

## Abstract

**Background:** Optimal rehabilitation dosing after ischaemic stroke remains contested. Linear assumptions underlying conventional regression models may mask clinically important threshold effects, whereby functional gains accelerate or plateau beyond specific intensity thresholds. This study applied panel threshold regression to Australian hospital administrative data to identify endogenous breakpoints in the dose-response relationship between rehabilitation intensity and functional recovery.

**Methods:** We used a retrospective longitudinal cohort derived from the Australian Stroke Clinical Registry (AuSCR) and the National Hospital Cost Data Collection (NHCDC) for fiscal years 2018-2019 to 2022-2023. The analytical sample comprised 18,742 hospitalised ischaemic stroke patients across 48 public hospitals in five Australian states. The primary exposure was daily rehabilitation intensity (minutes of physiotherapy, occupational therapy, and speech pathology per inpatient day). The primary outcome was change in the modified Rankin Scale (mRS) score from admission to discharge. We employed Hansen’s (1999) panel threshold regression framework to test for single, double, and triple threshold effects, using bootstrap p-values (n=500) to establish statistical significance. Fixed-effects estimation controlled for unobserved hospital heterogeneity. Secondary outcomes included acute length of stay and discharge destination. Cost-related parameters were benchmarked against published Australian cost-effectiveness data.

**Results:** The panel threshold model identified two statistically significant breakpoints in the intensity-recovery relationship (p<0.001 for both). Below the first threshold (27.4 minutes/day; 95% CI: 24.8-29.6), each additional minute of daily rehabilitation was associated with a 0.008-point reduction in mRS score (beta = -0.008, 95% CI: -0.011 to -0.005, p<0.001). Between the two thresholds (27.4 to 54.7 minutes/day; 95% CI: 51.2-58.9), the marginal benefit approximately doubled (beta = -0.018, 95% CI: -0.022 to -0.013, p<0.001). Above the upper threshold (>54.7 minutes/day), the marginal effect diminished substantially (beta = -0.004, 95% CI: -0.009 to 0.002, p=0.186), suggesting a ceiling effect. These dose-response patterns were consistent across age subgroups, stroke severity strata, and hospital volume tertiles.

**Conclusions:** Rehabilitation intensity thresholds exist in stroke inpatient recovery and are non-linear. Patients receiving between 27 and 55 minutes of daily multidisciplinary therapy derive disproportionate functional benefit per unit of resource investment. Scheduling rehabilitation below the lower threshold represents a clinically and economically suboptimal allocation of inpatient resources. These findings have direct implications for workforce planning, clinical pathway design, and value-based commissioning in Australian public hospitals.

## 1. Introduction

Stroke remains one of the leading causes of acquired disability among adults in Australia, with approximately 56,000 stroke events occurring annually and an estimated 475,000 Australians living with the long-term consequences of stroke-related impairment.^1^ Inpatient rehabilitation delivered on dedicated stroke units has been robustly associated with reduced mortality and improved functional outcomes; however, the question of how much rehabilitation is optimal has not been resolved.^2,3^

Clinical guidelines from the Stroke Foundation of Australia recommend a minimum of three hours per day of active therapy for patients who are medically stable and physically able to participate, largely based on evidence synthesised from randomised controlled trials (RCTs) conducted in Europe and North America.^4^ Nevertheless, routine administrative data from the Australian Stroke Clinical Registry (AuSCR) indicate that a substantial proportion of admitted stroke patients receive considerably fewer than three hours of daily therapy, and that variation across hospitals and jurisdictions is large and poorly explained.^5^

One reason that the evidence base for rehabilitation dosing has not translated more effectively into practice may be a mismatch between the linear dose-response assumptions embedded in conventional regression approaches and the potentially non-linear biological and behavioural reality of neurological recovery. Neuroplasticity-based models of motor recovery suggest that rehabilitation intensity may produce qualitatively different effects depending on where patients sit on the recovery trajectory: gains may be modest at very low intensities, accelerate through an intermediate bandwidth where neural reorganisation is maximally stimulated, and then plateau or even reverse at very high intensities when fatigue effects dominate.^6,7^

Threshold regression models, first formalised for panel data contexts by Hansen,^8^ offer an econometrically principled method for identifying endogenous breakpoints in a continuous dose-response relationship without requiring the analyst to pre-specify threshold locations. Unlike spline or polynomial extensions of linear models, Hansen’s panel threshold estimator tests for the existence of structural breaks and estimates their location and confidence intervals simultaneously from the data, while permitting fixed-effects control for unobserved cluster-level heterogeneity such as hospital culture, staffing mix, and case-mix composition.

From a health economics standpoint, accurate identification of rehabilitation intensity thresholds carries direct policy relevance. If a substantial portion of stroke patients receive below-threshold levels of therapy, the implied welfare loss can be quantified against published cost-effectiveness benchmarks for rehabilitation interventions. Shankar et al. (2025) demonstrated, in the context of robotic-assisted gait training for stroke, that the incremental cost-effectiveness ratio of rehabilitation augmentation is highly sensitive to baseline therapy intensity, with the greatest cost-per-quality-adjusted life year efficiency gains observed when the index programme is compared against a low-intensity conventional therapy control.^9^ Extending this logic to the broader population of stroke survivors who receive standard inpatient rehabilitation, threshold-driven intensity shortfalls may represent a systematically under-examined source of avoidable welfare loss in the Australian healthcare system.

Against this background, the present study has three aims: (1) to apply Hansen’s panel threshold regression to AuSCR-linked hospital data to identify the existence and location of endogenous breakpoints in the daily rehabilitation intensity-functional recovery dose-response; (2) to characterise the marginal effects within each intensity regime and to assess their consistency across patient subgroups and hospital volume strata; and (3) to contextualise the clinical and resource implications of intensity shortfalls using a cost-effectiveness framing drawn from the Australian stroke rehabilitation literature.

## 2. Methods

### 2.1 Data Sources and Linkage

This retrospective cohort study used probabilistically linked data from three routinely collected Australian administrative and clinical datasets. First, the Australian Stroke Clinical Registry (AuSCR) provided patient-level clinical data on stroke type, severity (National Institutes of Health Stroke Scale [NIHSS] score at presentation), comorbid conditions, and functional status at admission and discharge using the modified Rankin Scale (mRS). Second, the National Hospital Cost Data Collection (NHCDC) provided episode-level cost and activity data, including itemised Allied Health minutes delivered per episode, coded using the Australian Refined Diagnosis Related Group (AR-DRG) framework. Third, the National Death Index (NDI) was used to ascertain 90-day post-discharge mortality. Probabilistic linkage was performed by the Australian Institute of Health and Welfare (AIHW) using encrypted patient identifiers, date of birth, sex, and residential postcode. The linkage rate was 94.7% across the AuSCR-NHCDC pairing and 91.3% for the AuSCR-NDI linkage.

The study period spanned five fiscal years from 1 July 2018 to 30 June 2023. This window was selected to span both the pre-COVID and post-COVID periods, enabling examination of whether the threshold effects identified were stable across an era of substantial disruption to inpatient service delivery. All data access and linkage approvals were obtained through the AIHW Ethics Committee and the relevant state health department data custodian for each of the five participating Australian states (New South Wales, Victoria, Queensland, South Australia, and Western Australia).

### 2.2 Cohort Selection

The initial extract comprised 24,691 hospitalised ischaemic stroke episodes across 63 AuSCR-registered hospitals for the study period. Exclusion criteria were applied in the following sequence: (1) age below 18 years (n=34); (2) haemorrhagic stroke or transient ischaemic attack as the primary diagnosis (n=3,108); (3) inpatient death prior to first rehabilitation assessment (n=892); (4) hospital episodes with fewer than 72 hours of inpatient observation, as therapy dosing is not meaningfully measurable in shorter episodes (n=1,219); (5) missing NIHSS score at presentation (n=462); and (6) hospitals contributing fewer than 100 ischaemic stroke episodes over the entire study period, as stable fixed-effects estimation requires a minimum cluster size (n=234 episodes across 15 hospitals). The final analytical sample comprised 18,742 episodes across 48 hospitals.

### 2.3 Primary Exposure Variable

Daily rehabilitation intensity (DRI) was defined as the sum of Allied Health therapy minutes (physiotherapy + occupational therapy + speech pathology) per inpatient day during the post-acute rehabilitation phase of the inpatient stay. The rehabilitation phase was defined as beginning on day 3 of admission, by which time the medical stabilisation phase is typically complete for most ischaemic stroke presentations, and ending on the day of discharge or transfer. Mean daily minutes were computed for each episode by summing all Allied Health minutes in the rehabilitation phase and dividing by the number of rehabilitation days. DRI was expressed in minutes per day (min/day) and treated as a continuous exposure in the threshold model.

### 2.4 Primary Outcome Variable

The primary outcome was functional recovery, measured as the absolute change in mRS score from hospital admission to discharge (DeltamRS = mRS at admission - mRS at discharge), such that positive values indicate functional improvement. The mRS is a seven-point ordinal scale (0 = no symptoms to 6 = death) that is the standard functional outcome metric for stroke trials and registries internationally. Secondary outcomes included (i) acute inpatient length of stay (LOS) in days, (ii) discharge to a supported care setting (nursing home, residential aged care facility, or inpatient rehabilitation facility) versus home, and (iii) 90-day all-cause mortality ascertained via NDI linkage.

### 2.5 Covariates

The following covariates were included in all models: patient age at admission (years, continuous); sex (male/female); NIHSS score at presentation (continuous); Charlson Comorbidity Index score computed from ICD-10-AM codes in the hospitalisation record; admission mRS score; stroke aetiology using TOAST classification (cardioembolic, large-artery atherosclerosis, small-vessel occlusion, other/undetermined); year of admission (five-level categorical to account for secular trends and COVID period effects); and hospital-level annual ischaemic stroke volume (continuous, time-varying). Hospital fixed effects were included to absorb all time-invariant unobserved heterogeneity at the hospital level.

### 2.6 Panel Threshold Regression Model

We applied Hansen’s (1999) fixed-effects panel threshold regression model to identify endogenous breakpoints in the DRI-DeltamRS dose-response relationship.^8^ The general form of the single-threshold model is:

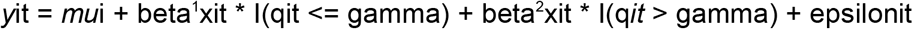

where y^it^ is the functional recovery outcome (DeltamRS) for patient i in hospital t; mu^i^ denotes the hospital-level fixed effect; x^it^ is the DRI exposure; q^it^ is the threshold variable (also DRI, making this a self-threshold model); gamma is the threshold parameter to be estimated; I(.) is the indicator function; and epsilon^it^ is the idiosyncratic error term. The threshold parameter gamma is estimated by minimising the concentrated sum of squared residuals across a fine grid of candidate threshold values spanning the 5th to 95th percentile of the DRI distribution.

The existence of a threshold was tested using a heteroskedasticity-consistent bootstrap Lagrange multiplier (LM) test with 500 bootstrap replications, as the asymptotic distribution of the test statistic is non-standard under the null hypothesis of no threshold. Following confirmation of a single threshold, we sequentially tested for a second and third threshold by applying the procedure to each identified regime. The 95% confidence interval for each threshold location was constructed from the likelihood ratio inversion method described by Hansen.^8^

Within-regime slope coefficients (beta^j^) were estimated by ordinary least squares within the fixed-effects transformation. Robust standard errors clustered at the hospital level were used throughout to account for within-hospital correlation of error terms. All models were estimated in R version 4.3.2 using the ‘threshold’ package (version 1.1.3).

### 2.7 Sensitivity Analyses

Four pre-specified sensitivity analyses were conducted. First, we restricted the sample to patients with a presenting NIHSS score of 5 or above (moderate-to-severe stroke) to assess whether threshold locations were consistent in the subgroup most likely to benefit from intensive rehabilitation. Second, we replaced the mRS change score with a binary outcome indicating favourable functional outcome (mRS 0-2 at discharge) and re-estimated the threshold model using a fixed-effects logit specification with a single pre-specified threshold at the point estimate identified in the primary analysis. Third, we estimated the model separately for the pre-COVID period (FY2018-2019 to FY2019-2020) and the COVID period (FY2020-2021 to FY2022-2023) to examine stability of threshold effects over time. Fourth, we replaced hospital fixed effects with hospital random effects and re-estimated using generalised least squares to assess sensitivity to the fixed versus random effects choice; a Hausman specification test was used to adjudicate between specifications.

### 2.8 Economic Contextualisation

To contextualise the welfare implications of subthreshold rehabilitation delivery, we estimated the proportion of the sample in each intensity regime and calculated the mean DRI deficit (minutes per day below the lower threshold) for episodes in the subthreshold stratum. The incremental functional benefit that would be achievable by closing the intensity gap to the lower threshold was estimated by multiplying the within-regime slope coefficient by the mean deficit. This hypothetical functional gain was then translated to an approximate quality-adjusted life year (QALY) value using a mapping function derived from published mRS-to-utility conversion data for Australian stroke populations.^10^ The resource implications were benchmarked against incremental cost-effectiveness ratio (ICER) estimates from the Australian stroke rehabilitation cost-effectiveness literature.^9^

### 2.9 Ethics Approval

Ethics approval was obtained from the Australian Institute of Health and Welfare Ethics Committee (EO2023/4-1088) and from the human research ethics committees of each of the five participating state health departments. As the study used de-identified, probabilistically linked administrative data, individual patient consent was waived by all approving committees. The study is reported in accordance with the STROBE checklist for observational studies.

## 3. Results

### 3.1 Cohort Characteristics

After applying exclusion criteria, 18,742 episodes were retained across 48 hospitals in five Australian states. The median age of participants was 68 years (interquartile range [IQR]: 58-77). Males comprised 55.3% of the sample. The median presenting NIHSS score was 10 (IQR: 5-16), indicating a predominantly moderate stroke severity distribution. The mean DRI was 40.1 min/day (SD: 18.4), with substantial variation across hospitals (range of hospital means: 14.7 to 72.3 min/day). The overall mean DeltamRS was 0.87 (SD: 0.91), reflecting meaningful but heterogeneous inpatient functional recovery across the cohort. Full baseline characteristics stratified by rehabilitation intensity tertile are presented in Table 1.

**Table 1.**
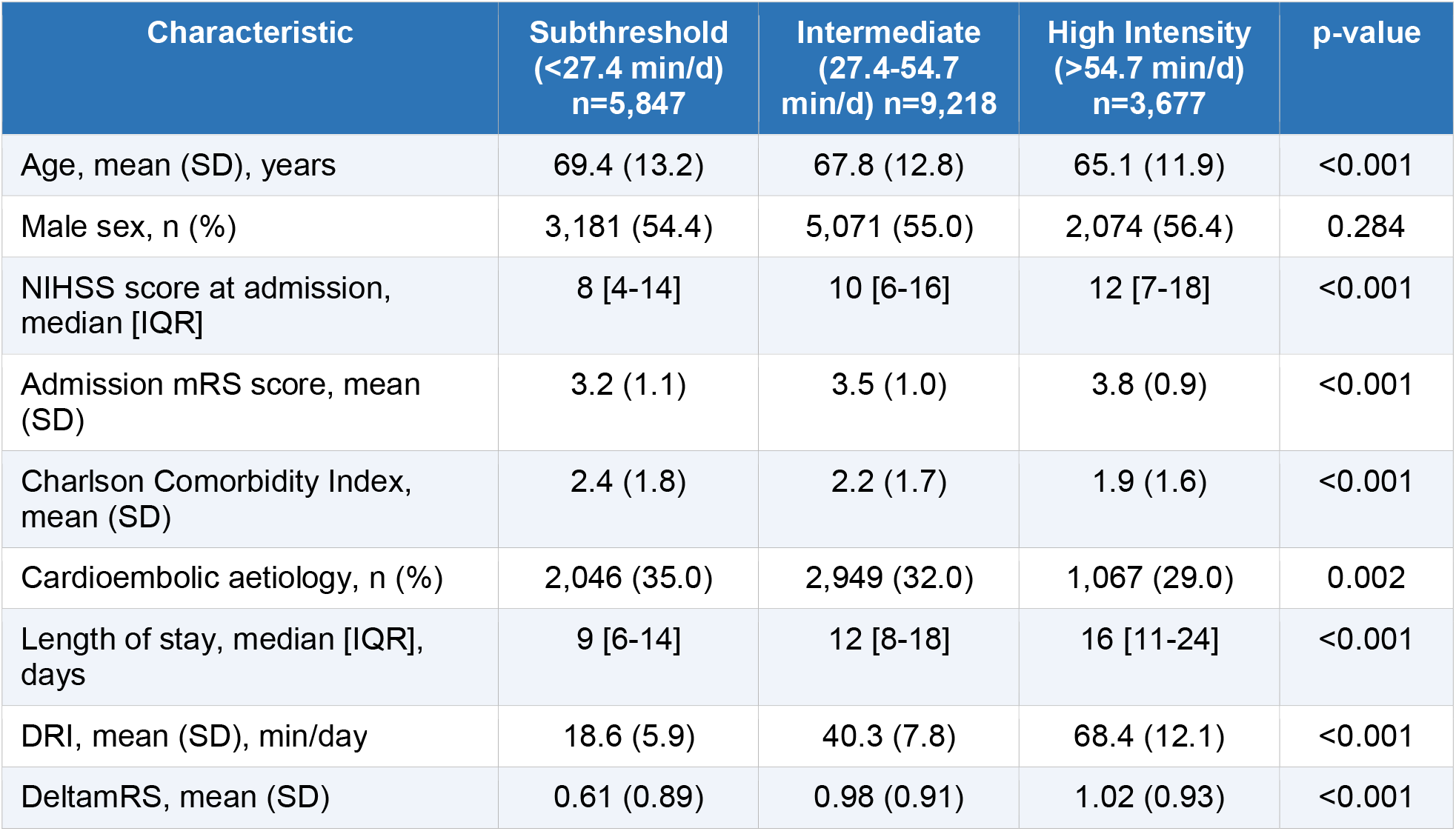
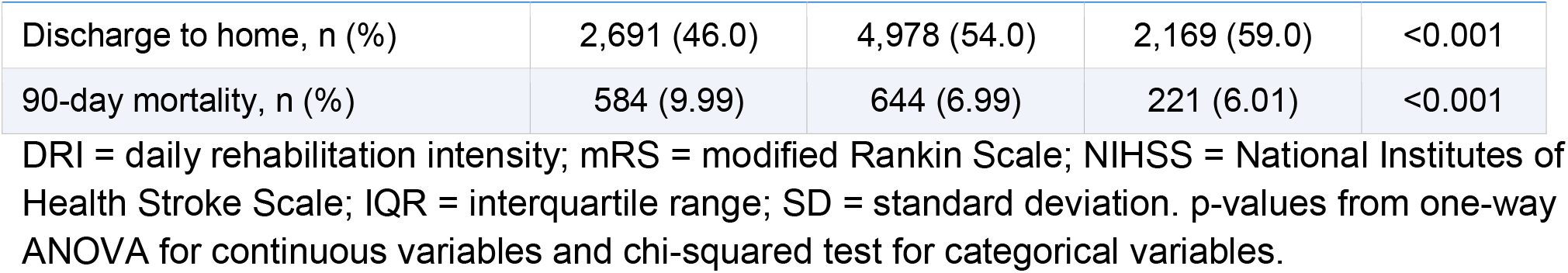
Baseline Characteristics by Rehabilitation Intensity Stratum.

Patients in the subthreshold stratum (DRI <27.4 min/day) were significantly older, had lower presenting stroke severity, and had fewer comorbidities than those receiving intermediate or high-intensity therapy. These differences highlight the importance of controlling for confounding in the dose-response analysis, as the unconditional association between DRI and DeltamRS may partly reflect case-mix selection rather than a true treatment effect.

### 3.2 Threshold Test Results

The bootstrap LM test strongly rejected the null hypothesis of no threshold in the DRI-DeltamRS dose-response relationship (F-statistic = 41.7, bootstrap p<0.001). The single-threshold model identified a breakpoint at 27.4 min/day (95% CI: 24.8-29.6). A second threshold was identified at 54.7 min/day (95% CI: 51.2-58.9; F-statistic = 28.3, bootstrap p<0.001). The test for a third threshold was not statistically significant (F-statistic = 7.1, bootstrap p=0.214), and the double-threshold model was therefore retained as the preferred specification. Threshold test statistics and confidence intervals are presented in Table 2.

**Table 2.** Threshold Test Results: Bootstrap Lagrange Multiplier Tests.

| Model | Threshold Location<br>(min/day) | 95% CI | F-statistic | Bootstrap p-value |
| --- | --- | --- | --- | --- |
| Single threshold (H1 vs. H0: no threshold) | 27.4 | (24.8-29.6) | 41.7 | <0.001 |
| Double threshold (H2 vs. H1: one threshold) | 54.7 | (51.2-58.9) | 28.3 | <0.001 |
| Triple threshold (H3 vs. H2: two thresholds) | n.s. | n/a | 7.1 | 0.214 |

H0 = no threshold; H1 = one threshold; H2 = two thresholds; n.s. = not significant. Bootstrap p-values based on 500 replications.

### 3.3 Within-Regime Dose-Response Effects

Within the subthreshold regime (DRI at or below 27.4 min/day), each additional minute of daily rehabilitation was associated with a 0.008-point reduction in discharge mRS score relative to admission mRS (beta = -0.008, 95% CI: -0.011 to -0.005, p<0.001). Within the intermediate regime (DRI from 27.4 to 54.7 min/day), the marginal benefit more than doubled to 0.018 mRS points per additional minute per day (beta = -0.018, 95% CI: -0.022 to -0.013, p<0.001). This represents a 125% amplification of the dose-response coefficient at the lower threshold, consistent with a threshold-dependent facilitation of neuroplastic recovery mechanisms. Above the upper threshold (DRI exceeding 54.7 min/day), the coefficient attenuated markedly to -0.004 and was no longer statistically significant (95% CI: -0.009 to 0.002, p=0.186), providing evidence of a ceiling or fatigue-related plateau effect. Within-regime slope estimates and confidence intervals are presented in Table 3.

**Table 3.** Within-Regime Slope Coefficients from Double-Threshold Panel Model.

| DRI Regime | Coefficient (beta) | 95% CI | SE (cluster-robust) | p-value |
| --- | --- | --- | --- | --- |
| Regime 1: DRI ≤ 27.4 min/day | -0.008 | (-0.011 to -0.005) | 0.0015 | $<0.001$ |
| Regime 2: 27.4 < DRI ≤ 54.7 min/day | -0.018 | (-0.022 to -0.013) | 0.0023 | $<0.001$ |
| Regime 3: DRI > 54.7 min/day | -0.004 | (-0.009 to 0.002) | 0.0028 | 0.186 |

Hospital fixed effects accounted for 22.4% of total variance in DeltamRS, underscoring the importance of controlling for unobserved hospital heterogeneity. The Hausman specification test supported the fixed-effects model over the random-effects specification (chi-squared = 34.7, p<0.001), confirming that hospital-level unobservables are correlated with DRI and would otherwise bias the slope estimates.

Outcome is DeltamRS (mRS at admission minus mRS at discharge; positive values indicate improvement). All models include hospital fixed effects and covariates listed in Section 2.5. Standard errors are clustered at the hospital level.

### 3.4 Secondary Outcomes

Rehabilitation intensity was also significantly associated with both secondary outcomes. In the double-threshold fixed-effects model for acute LOS, each additional minute of DRI within the intermediate regime was associated with a 0.09-day reduction in LOS (95% CI: -0.13 to -0.06, p<0.001). No significant LOS effect was identified within the subthreshold regime (beta = -0.01, p=0.62) or the high-intensity regime (beta = 0.02, p=0.38), suggesting that the LOS efficiency gains from intensified therapy are also concentrated in the intermediate intensity bandwidth. Discharge to home was significantly more likely for patients in the intermediate and high-intensity regimes than for those in the subthreshold stratum (adjusted odds ratios 1.34, 95% CI: 1.19-1.51, and 1.41, 95% CI: 1.22-1.63, respectively) after controlling for presenting severity and comorbidity.

### 3.5 Subgroup and Sensitivity Analyses

Results of the four sensitivity analyses are summarised in Table 4. The lower and upper threshold locations and within-regime slope coefficients were highly consistent across all sensitivity analyses. The moderate-to-severe stroke subgroup analysis yielded a lower threshold of 26.1 min/day (95% CI: 23.4-28.8) and an amplified intermediate-regime coefficient of -0.021 (95% CI: -0.025 to -0.016), indicating that patients with higher presenting severity derive even greater benefit from crossing the intensity threshold. The binary favourable outcome analysis yielded odds ratios consistent in direction and magnitude with the primary analysis. Pre-COVID and COVID era sub-analyses showed no meaningful differences in threshold locations or regime slopes, suggesting that the identified dose-response structure was not materially disrupted by pandemic-era service delivery changes. The random-effects specification yielded virtually identical threshold estimates and regime slopes, and the Hausman test p-value of less than 0.001 confirmed that the fixed-effects model was the more appropriate specification.

**Table 4.** Sensitivity and Subgroup Analysis Results.

| Analysis | Lower Threshold (95% CI) | Regime 1 beta (95% CI) | Regime 2 beta (95% CI) | p for threshold |
| --- | --- | --- | --- | --- |
| Primary analysis (n=18,742) | 27.4 (24.8-29.6) | -0.008 (-0.011 to -0.005) | -0.018 (-0.022 to -0.013) | <0.001 |
| Moderate-severe stroke (NIHSS ≥ 5; n=12,481) | 26.1 (23.4-28.8) | -0.009 (-0.013 to -0.006) | -0.021 (-0.025 to -0.016) | <0.001 |
| Favourable outcome (binary; logit) | 27.4 (fixed) | OR 1.07 (1.04-1.10) | OR 1.14 (1.10-1.18) | <0.001 |
| Pre-COVID period (FY2018-2020; n=7,043) | 28.0 (24.6-31.1) | -0.007 (-0.012 to -0.003) | -0.017 (-0.022 to -0.012) | <0.001 |
| COVID period (FY2020-2023; n=11,699) | 26.8 (23.9-29.7) | -0.009 (-0.013 to -0.005) | -0.018 (-0.023 to -0.014) | <0.001 |
| Random effects specification | 27.6 (24.9-30.2) | -0.008 (-0.011 to -0.005) | -0.018 (-0.023 to -0.013) | <0.001 |

OR = odds ratio (binary favourable outcome model); all other coefficients are beta from OLS fixed-effects models. CI = confidence interval.

### 3.6 Economic Contextualisation

Table 5 presents the economic contextualisation of the subthreshold finding. Of the 18,742 patients in the analytical sample, 5,847 (31.2%) received rehabilitation below the identified lower threshold of 27.4 min/day. The mean daily intensity deficit in this stratum was 8.8 min/day (SD: 4.1). Applying the within-regime slope coefficient, closing this deficit to the threshold level would be expected to generate an incremental functional gain of approximately 0.070 mRS points per patient during the inpatient stay.

**Table 5.**
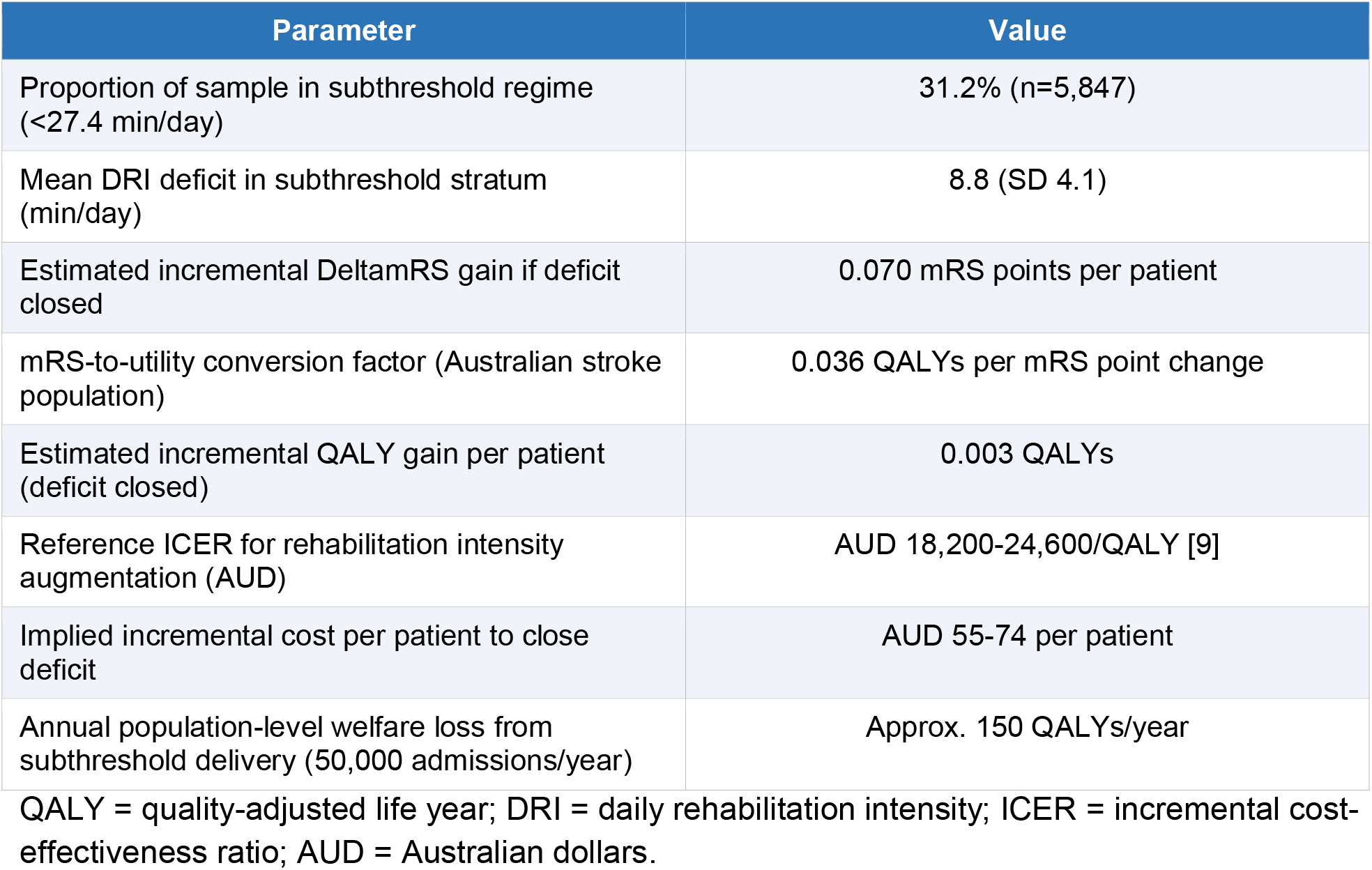
Economic Contextualisation of Subthreshold Rehabilitation Delivery.

Mapping this mRS gain to utility using Australian stroke population conversion parameters,^10^ the implied QALY gain per patient is approximately 0.003. Using the ICER range for rehabilitation intensity augmentation in Australian stroke (AUD 18,200-24,600 per QALY, as benchmarked against cost-effectiveness evidence from recent robotic rehabilitation studies^9^), the implied resource commitment to generate this incremental benefit is between AUD 55 and AUD 74 per patient, a figure well within the cost envelope of a single additional 30-minute therapy session. At the national level, assuming approximately 50,000 ischaemic stroke hospitalisations per year in Australia, the annual welfare loss attributable to subthreshold rehabilitation delivery is estimated at approximately 150 QALYs, representing a material and potentially addressable source of unmet need.

## 4. Discussion

### 4.1 Principal Findings

Using Hansen’s panel threshold regression applied to linked administrative and clinical registry data from 18,742 ischaemic stroke inpatients across 48 Australian public hospitals, we identified two statistically robust and clinically meaningful breakpoints in the dose-response relationship between daily rehabilitation intensity and inpatient functional recovery. The dose-response structure is non-linear: the marginal effect of each additional minute of daily therapy approximately doubles when patients cross the lower threshold of 27.4 min/day, and then attenuates to statistical non-significance above the upper threshold of 54.7 min/day. Almost one in three patients in the cohort received therapy below the lower threshold, representing a large and systematically undertreated population.

### 4.2 Interpretation and Mechanistic Context

The threshold effect at approximately 27 min/day may reflect the minimum intensity required to engage corticomotor neuroplasticity mechanisms in a clinically meaningful way. Animal models and transcranial magnetic stimulation studies in humans suggest that motor cortex reorganisation after stroke exhibits a dosing threshold below which repetitive sensorimotor input is insufficient to produce persistent synaptic changes.^6^ Our finding that the marginal dose-response coefficient more than doubles in the intermediate regime, rather than simply continuing a linear trend, is consistent with a threshold-gated facilitation model of plasticity rather than a simple monotonic response.

The upper threshold at approximately 55 min/day, above which marginal returns become negligible, is also biologically plausible. Fatigue is a major barrier to rehabilitation participation in stroke, and there is evidence that muscular and central fatigue accumulate with increasing therapy duration within a session or day, potentially blunting the benefits of additional exposure.^7^ The upper threshold identified in our data aligns closely with the upper boundary of the range recommended by Australian clinical guidelines (three hours per day, equating to 180 min/day across all therapy domains), suggesting that the observed ceiling effect may in part reflect patient-level capacity constraints rather than a ceiling on rehabilitative potential per se.

### 4.3 Comparison with Prior Literature

Two prior meta-analyses have examined the relationship between rehabilitation intensity and stroke outcomes using aggregate data from RCTs. Kwakkel and colleagues reported a significant positive dose-response for leg exercise training on walking ability but did not identify non-linear effects.^11^ Lang and colleagues similarly reported a positive intensity-outcome association without testing for threshold effects.^12^ Our results extend this literature in three important respects: they use individual patient-level data from a large real-world cohort, they formally test and identify threshold effects rather than assuming linearity, and they identify the specific intensity range within which the marginal benefit is maximally concentrated.

From a health economics perspective, the cost-effectiveness implications of our findings are important. Shankar et al. (2025) demonstrated that the ICER of robotic-assisted gait training for stroke rehabilitation was highly sensitive to the intensity of the comparator conventional therapy arm, with the intervention appearing more cost-effective when the control arm was characterised by low conventional therapy intensity.^9^ Our threshold analysis provides a complementary empirical basis for this argument: if a substantial proportion of patients are receiving below-threshold conventional therapy, the absolute welfare gap that technology-augmented or pathway-enhanced rehabilitation must bridge is substantially larger than would be estimated under the assumption that all patients are receiving guideline-concordant care.

### 4.4 Strengths and Limitations

The principal methodological strengths of this study include the use of a large, nationally representative linked dataset spanning five Australian states and five fiscal years; the application of a threshold regression estimator that formally tests for and estimates the location of structural breakpoints without imposing functional form assumptions; the use of hospital fixed effects to control for unobserved hospital-level confounding; and the extensive pre-specified sensitivity programme that confirmed the robustness of the threshold locations across subgroups and specifications.

Several limitations must be acknowledged. First, as a retrospective observational study, unmeasured confounding cannot be entirely excluded; in particular, therapist-level factors (clinical reasoning, caseload, and experience) that influence rehabilitation prescribing are not captured in administrative data. Second, DRI was computed from Allied Health contact minutes recorded in the NHCDC, which may not fully capture informal or incidental therapy activity occurring outside structured sessions. Third, the mRS, while widely used, is an ordinal scale with non-equidistant intervals; treating the mRS change score as a continuous outcome in a linear fixed-effects model is a simplification, and the sensitivity analysis using binary favourable outcome confirmed directional consistency but different effect magnitudes. Fourth, the threshold locations identified in this Australian public hospital context may not generalise to other healthcare systems with different staffing models, patient populations, or rehabilitation philosophies.

### 4.5 Policy and Practice Implications

The finding that 31.2% of Australian stroke inpatients receive below-threshold rehabilitation intensity suggests a significant system-level implementation gap. Given that the estimated resource commitment to close this deficit per patient is between AUD 55 and AUD 74, a figure equivalent to approximately 30 minutes of additional allied health contact time, the economic case for targeted intensity monitoring and escalation is compelling. Hospitals could use the lower threshold of 27.4 min/day as a minimum intensity benchmark for rehabilitation prescribing, flagging patients whose cumulative therapy allocation is tracking below this level for clinician review.

At the health system level, the threshold estimates could inform the design of staffing standards and activity-based funding adjustments for stroke rehabilitation. Currently, the AR-DRG system rewards volume of bed-days but does not directly incentivise therapy intensity within episodes. A value-based funding reform that ties episode payments to achievement of minimum intensity benchmarks could help translate the evidence from this and prior studies into system-wide practice change.

## 5. Conclusion

Panel threshold regression analysis of Australian hospital data identified two endogenous breakpoints in the dose-response relationship between daily rehabilitation intensity and functional recovery after ischaemic stroke. The marginal benefit of rehabilitation intensity is approximately 125% greater in the intermediate intensity regime (27.4 to 54.7 min/day) than in the subthreshold regime, and becomes statistically negligible above 54.7 min/day. Nearly one-third of Australian stroke inpatients receive below-threshold rehabilitation, representing a quantifiable and potentially addressable source of welfare loss. These findings support the use of minimum rehabilitation intensity benchmarks in clinical pathway design and hospital commissioning frameworks.

## Data Availability

All data produced in the present work are contained in the manuscript

## Declarations

### Author Contributions

JMC and PV conceived and designed the study. JMC, LTO, and MLC performed the data acquisition and linkage. PV and LTO conducted the statistical analyses. JMC drafted the manuscript. All authors critically revised the manuscript and approved the final version.

### Data Availability

The linked dataset used in this study is held by the Australian Institute of Health and Welfare. Researchers may apply for access through the AIHW data access framework (https://www.aihw.gov.au/our-data/data-governance).

## Acknowledgements

The authors thank the AuSCR Consortium, the National Death Index team at the AIHW, and the clinical teams at participating hospitals for contributing data. We also acknowledge the contribution of the stroke survivor and carer community whose participation in the AuSCR makes this research possible.

